# First-Trimester Multi-modal cfDNA Analysis for Prediction of Preterm and Term Preeclampsia

**DOI:** 10.64898/2026.03.12.26348234

**Authors:** Rebecca Ertl, Argyro Syngelaki, Olga Frank, Lukas Lüftinger, Eva Lukáčová, Casper Lumby, Adrian Stütz, Stephan Beisken, Andreas E. Posch, Kypros H. Nicolaides

## Abstract

**Background:** Preeclampsia, which is a leading cause of maternal and perinatal mortality and morbidity, represents a biologically heterogeneous syndrome. First-trimester screening with the Fetal Medicine Foundation competing-risks model enables prevention of preterm preeclampsia through aspirin prophylaxis but depends on Doppler velocimetry and biochemical measurements that limit scalability and offer limited discrimination for term disease. A unified, molecular first trimester test capable of stratifying risk across the full clinical spectrum of preeclampsia has not been established.

**Objective:** To determine whether multi-modal, tissue-resolved analysis of first trimester circulating cell-free DNA (cfDNA), obtained during routine non-invasive prenatal testing (NIPT), enables early prediction of both preterm and term preeclampsia.

**Study Design:** This nested case-control study included 125 singleton pregnancies sampled at 11–14 weeks’ gestation after quality control (48 controls, 30 preterm preeclampsia, 47 term preeclampsia). For 80 pregnancies, matched placental villi and maternal buffy coat samples were available to derive tissue reference profiles. Plasma cfDNA underwent multi-modal sequencing using Oxford Nanopore Technologies, enabling tissue-resolved analysis of fragmentomic and epigenetic signatures. Separate ensemble machine-learning classifiers were developed for preterm (<37 weeks) and term (≥37 weeks) preeclampsia using stratified 10-fold cross-validation. Model discrimination was evaluated using area under the receiver operating characteristic curve (AUROC), sensitivity at predefined specificity thresholds, and comparison with the FMF first-trimester risk score. A population-level simulation of 100,000 pregnancies, applying incidence point estimates of 2.5% for preterm and 7.5% for term PE, was used to derive predictive values and likelihood ratios.

**Results:** The multi-modal cfDNA classifier achieved an AUROC (95% CI) of 0.85 (0.77–0.91) for preterm preeclampsia and 0.84 (0.76–0.91) for term preeclampsia. The FMF score yielded an AUROC of 0.80 (0.70–0.89) for preterm and 0.53 (0.43–0.63) for term PE. At 80% specificity, cfDNA sensitivity was 70.5% for preterm and 72.1% for term preeclampsia, demonstrating improved discrimination for term disease compared with FMF screening. In simulated population-level analysis, positive likelihood ratios were 4.25 (preterm) and 3.83 (term), with negative likelihood ratios of 0.21 and 0.34, respectively, supporting meaningful post-test risk stratification and strong rule-out performance.

**Conclusion:** First-trimester multi-modal, tissue-resolved cfDNA analysis enables early risk stratification across the full clinical spectrum of preeclampsia from a single routine blood sample. Compared with FMF screening, this approach can potentially improve discrimination for term preeclampsia while providing incremental improvement for preterm disease. The potential for integration into existing NIPT workflows offers a scalable pathway toward unified precision prevention, supporting timely aspirin prophylaxis for preterm preeclampsia and risk-adapted surveillance strategies for term disease.

## Introduction

Preeclampsia is a pregnancy-specific syndrome with complex causes, mechanisms, prediction, management, and prevention challenges.^1^ It remains a leading cause of maternal and perinatal morbidity and mortality worldwide.^2^ Short-term U.S. healthcare costs attributable to preeclampsia were estimated at $2.18 billion in 2012 at 3.8% incidence.^3^ With incidence rising to 7.9% nation-wide in 2021 as well as exceeding 12% in recent multicenter data, and after adjustment for medical cost inflation, the projected annual short-term burden exceeds $7–10 billion by 2026 in the United States alone.^4–6^ Beyond this immediate impact, preeclampsia results in a two- to four-fold increase in lifetime cardiometabolic disease risk among affected women and increased vulnerability to prematurity-related, later cardiovascular and neurodevelopmental disorders in their offspring.^1,7^

Reducing this burden requires effective early risk stratification and prevention. Randomized trials and meta-analyses demonstrate that low-dose aspirin initiated before 16 weeks’ gestation substantially reduces preterm preeclampsia, whereas later initiation provides limited benefit.^8,9^ The Fetal Medicine Foundation (FMF) first-trimester algorithm, combining maternal characteristics, mean arterial pressure, uterine artery Doppler, and serum placental growth factor or pregnancy associated plasma protein-A, achieves robust detection of preterm preeclampsia.^10,11^ However, performance for term preeclampsia is modest, and reliance on Doppler velocimetry and specialized assays limits scalability in many countries. Although third-trimester angiogenic biomarkers, particularly the soluble Fms-like tyrosine kinase-1-to-placental growth factor (sFlt-1/PlGF) ratio, improve short-term prediction of severe disease, universal late-pregnancy testing has not been adopted because it is operationally and economically infeasible without earlier risk enrichment.^12^

Preeclampsia is a heterogeneous syndrome in which diverse etiologic pathways converge on endothelial dysfunction, inflammation, and syncytiotrophoblast stress.^1,13^ Placental ischemia and angiogenic imbalance predominate in preterm disease, whereas term preeclampsia comprises subgroups with and without an antiangiogenic profile that differ in inflammatory signatures and clinical trajectories.^14^ This heterogeneity likely contributes to the uneven performance of current screening strategies.

Circulating cell-free DNA (cfDNA), obtained through routine non-invasive prenatal testing, offers a scalable molecular platform for early risk assessment.^15–17^ Beyond fetal aneuploidy detection, cfDNA contains fragmentomic and epigenetic tissue-of-origin signals reflective of placental dysfunction and maternal vascular stress. Prior studies, however, have largely focused on single molecular modalities and preterm preeclampsia, with limited applicability to term disease.^18–22^

Whether multi-modal, tissue-resolved first-trimester cfDNA analysis can enable unified risk stratification across the full preeclampsia spectrum remains unknown. We evaluate a cfDNA framework for early prediction of both preterm and term preeclampsia (Figure 1). A single first-trimester blood test could operationalize the “turning the pyramid of care” paradigm, shifting care from reactive management to proactive, personalized prevention and surveillance of maternal and fetal risk.^23^

**Figure 1:**
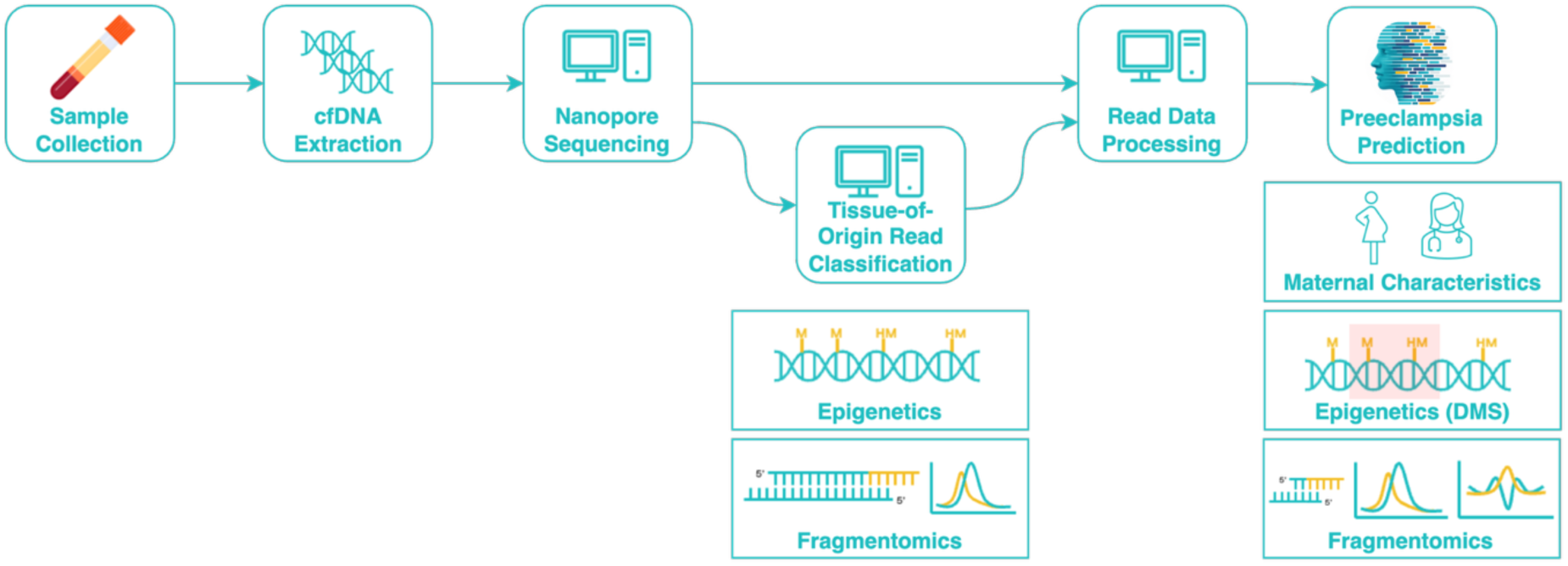
Schematic overview of the analytical framework evaluated in this study. A tissue-of-origin read classifier integrates epigenetic and fragmentomic features at the individual cfDNA read level to infer placental and maternal tissue contributions. The tissue-resolved preeclampsia prediction model combines maternal characteristics, differentially methylated CpG segments (DMS), and fragmentomic features to generate risk estimates for preterm and term preeclampsia.

## Material and Methods

### Study Design

This study was conducted as a nested case-control analysis within a pregnancy cohort recruited at King’s College Hospital, London, UK. Biobanked samples from women with singleton pregnancies were selected if a routine first-trimester hospital visit had been performed, and complete data on pregnancy outcome, maternal characteristics, medical history, and clinical measurements were available. Pregnancies with multifetal gestation, major fetal structural anomalies, or fetal aneuploidy were excluded.

Cases were defined as pregnancies complicated by preeclampsia, diagnosed according to the criteria of the American College of Obstetricians and Gynecologists. In preeclampsia there is new-onset hypertension after 20 weeks’ gestation in previously normotensive women, defined as systolic blood pressure ≥140 mmHg and/or diastolic blood pressure ≥90 mmHg on at least two occasions at least 4 hours apart, together with proteinuria (≥300 mg in a 24-hour urine collection, urinary protein–creatinine ratio ≥30 mg/mmol, or ≥2+ on dipstick testing), or – in the absence of proteinuria – new-onset hypertension with systemic features including thrombocytopenia, renal insufficiency, impaired liver function, pulmonary oedema, or cerebral or visual symptoms.^24^ Cases were stratified by gestational age at delivery into preterm (<37+0 weeks) and term preeclampsia (≥37+0 weeks). Control pregnancies were defined as those without hypertensive pregnancy disorders and delivery at ≥37+0 weeks.

This study was conducted in accordance with the ethical standards for human research established by the Declaration of Helsinki. The original data collection received approval from the London-Surrey Borders Research Ethics Committee. Written informed consent was obtained from all participants prior to inclusion in the study.

### Sample Collection and Processing

Maternal peripheral blood was collected at the routine first-trimester hospital visit. Blood was drawn into EDTA tubes and processed according to a standardized protocol. Plasma and, for a subset of samples, buffy coat were separated within 30 minutes of collection, aliquoted, and stored at −80 °C until further analysis.

For the same subset, placental tissue samples were collected by chorionic villus sampling. All tissue samples were processed and stored using standardized procedures to preserve DNA integrity.

Clinical data included maternal demographic characteristics, obstetric and medical history, and clinical measurements obtained at the first-trimester visit. Pregnancy outcomes, including gestational age at delivery, birth weight, and diagnosis of preeclampsia, were recorded after delivery.

The final cohort included 133 pregnancies: 52 controls, 50 term, and 31 preterm preeclampsia; matched placental and maternal buffy coat samples were available for 80 pregnancies and processed to generate reference fragmentomic and epigenetic profiles. Samples failing quality control due to identity inconsistencies, genomic DNA contamination or degradation, or insufficient sequencing output were excluded. High-quality cfDNA and reference tissue datasets were retained for downstream fragmentomic, methylation, and risk prediction analyses (Figure 2).

**Figure 2:**
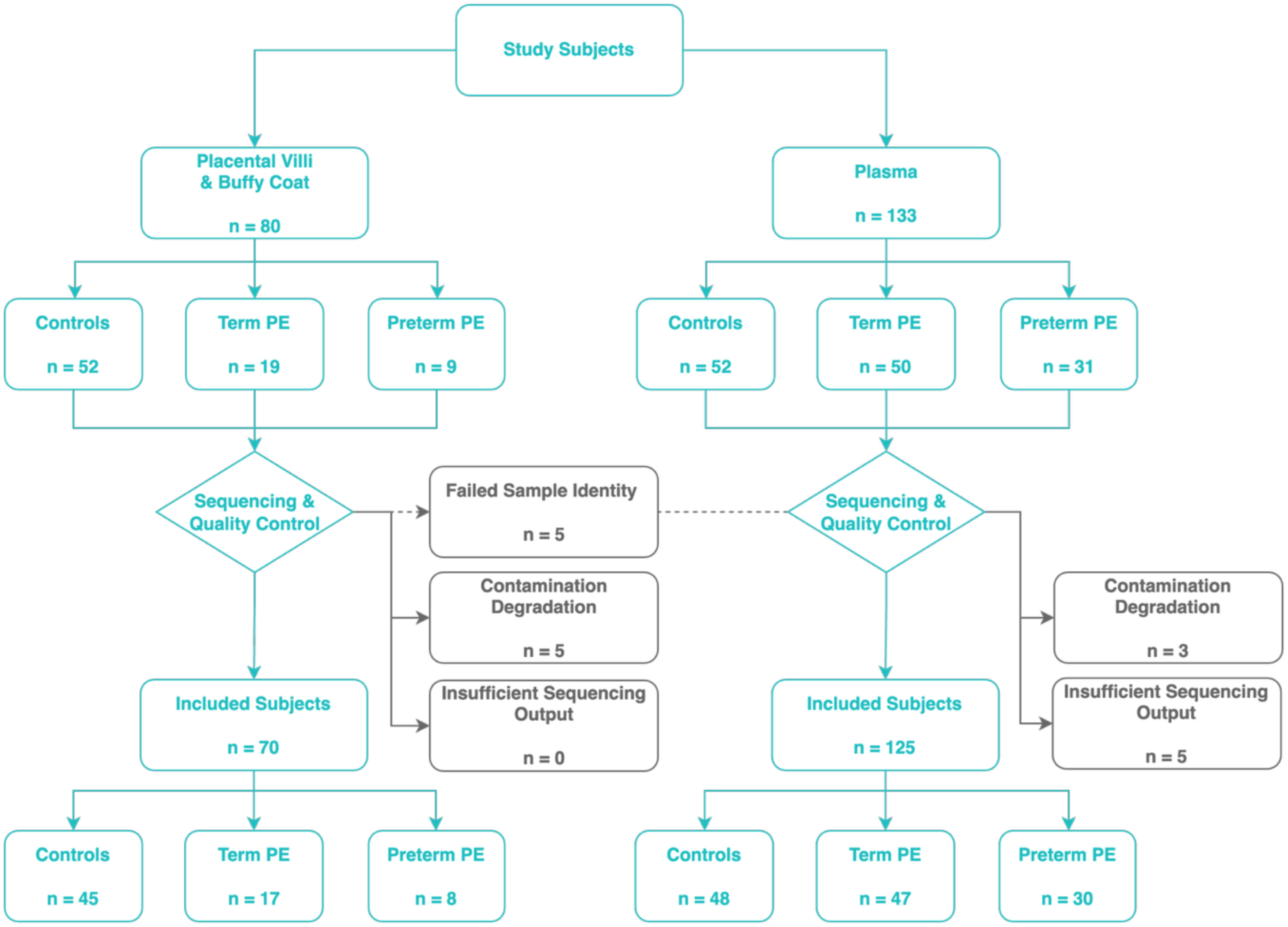
Study flow-diagram. Maternal plasma samples from 133 pregnancies (52 controls, 50 term preeclampsia, and 31 preterm preeclampsia) were processed for cfDNA extraction, sequencing, and bioinformatic analysis. For a subset of 80 pregnancies, matched maternal buffy coat and placental villi samples were additionally sequenced. All samples underwent standardized quality control, and samples failing quality thresholds were excluded prior to downstream analyses.

### DNA Extraction, Sequencing and Processing

cfDNA was extracted from maternal plasma using the aitios® cfDNA standardized protocol v1 – a validated extraction workflow preserving native cfDNA fragment length distributions – and sequenced on an Oxford Nanopore Technologies (ONT) platform (supplementary data: DNA Extraction and Sequencing). Sequencing data derived from both cfDNA and tissue gDNA samples were processed using a unified bioinformatics pipeline (supplementary data: Bioinformatics Processing).

### Multi-Modal Preeclampsia Prediction

The multi-modal, tissue-resolved preeclampsia classifier inferred risk for preterm and term disease from cfDNA fragmentomic and epigenomic signals together with maternal characteristics (Supplementary Data: Multi-Modal Preeclampsia Prediction). Separate classifiers were trained for preterm and term preeclampsia to enable disease subtyping, with pregnancies affected by the respective subtype as the positive class and all remaining samples as controls. Model training followed a modular ensemble framework (Supplementary Data: Model Development).

### Model Evaluation

Model performance was assessed using stratified 10-fold cross-validation. Performance was quantified using the area under the receiver operating characteristic (AUROC) curve. Sensitivity and accuracy were reported at specificity thresholds of 70%, 80%, and 90% from cross-validated model scores.

For benchmarking, ROC curves were constructed using the FMF first-trimester risk score. AUROCs of the cfDNA-based models and the FMF risk score were compared for preterm and term preeclampsia using DeLong’s test for correlated ROC curves. ROC curves were also generated for a combined model including cfDNA features (including maternal characteristics) and the FMF risk score to assess complementary predictive value.

To estimate clinically relevant predictive values, we performed a population-level screening simulation. The training dataset was bootstrapped to 100,000 pregnancies assuming prevalences of 2.5% for preterm, 7.5% for term, and 10% overall preeclampsia, reflecting recent U.S. incidence estimates from a prospective multi-center study. Confusion matrices at predefined specificity thresholds were used to calculate predictive values, likelihood ratios, post-test probabilities, and post-test odds.

To explore the biological processes captured by predictive cfDNA models, functional enrichment analysis was performed in plasma cfDNA methylation changes (supplementary data: Functional Enrichment Analysis).

## Results

### Cohort Characterization

Maternal plasma samples were obtained from 133 singleton pregnancies at 11+0 – 13+6 weeks’ gestation. Eight plasma cfDNA samples were excluded after quality control (two genomic DNA contamination, one genetic discordance with matched buffy coat, and five insufficient sequencing yield), leaving 125 plasma samples: 48 controls, 30 preterm preeclampsia, and 47 term preeclampsia (Table 1).

**Table 1:**
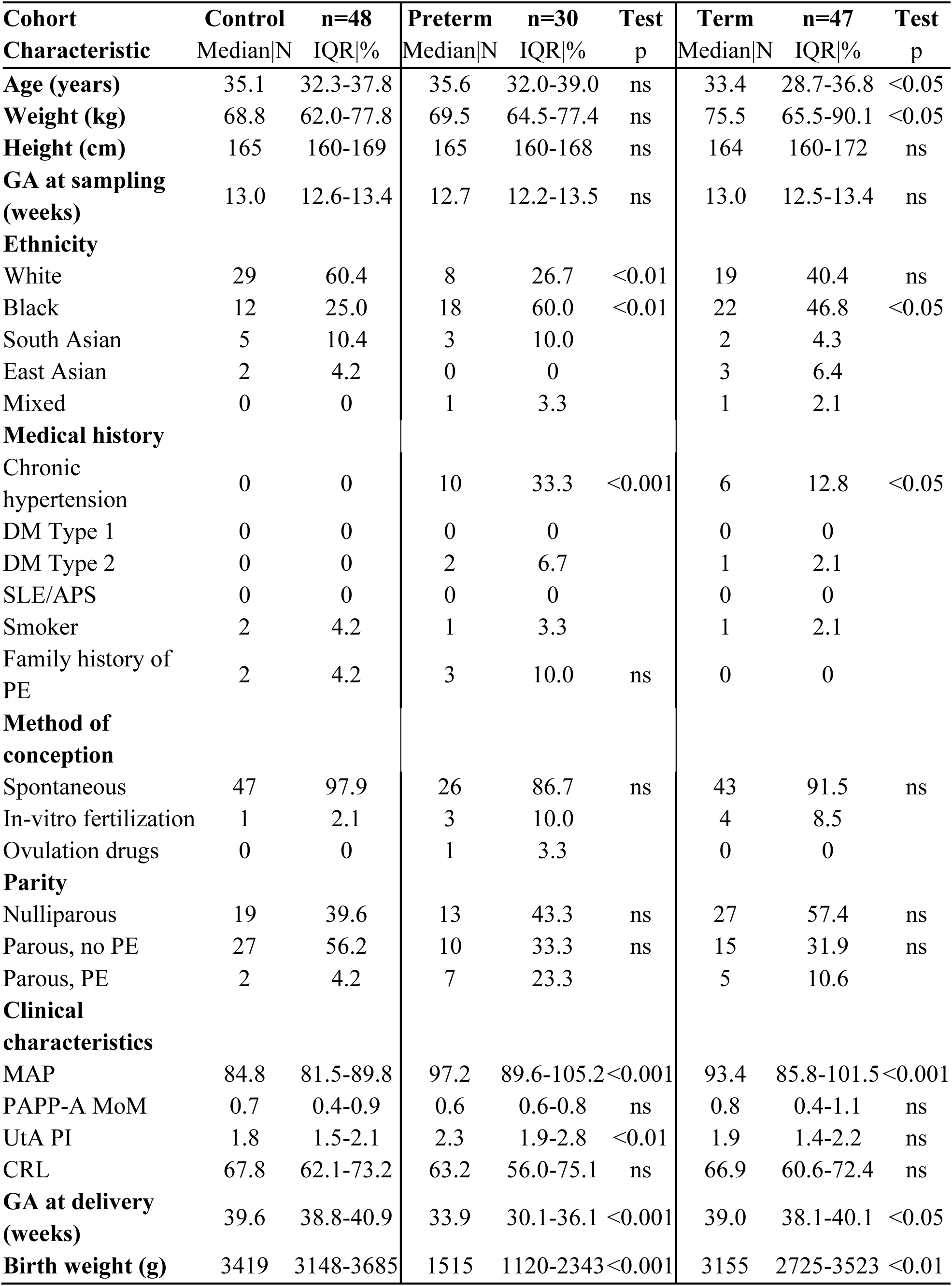
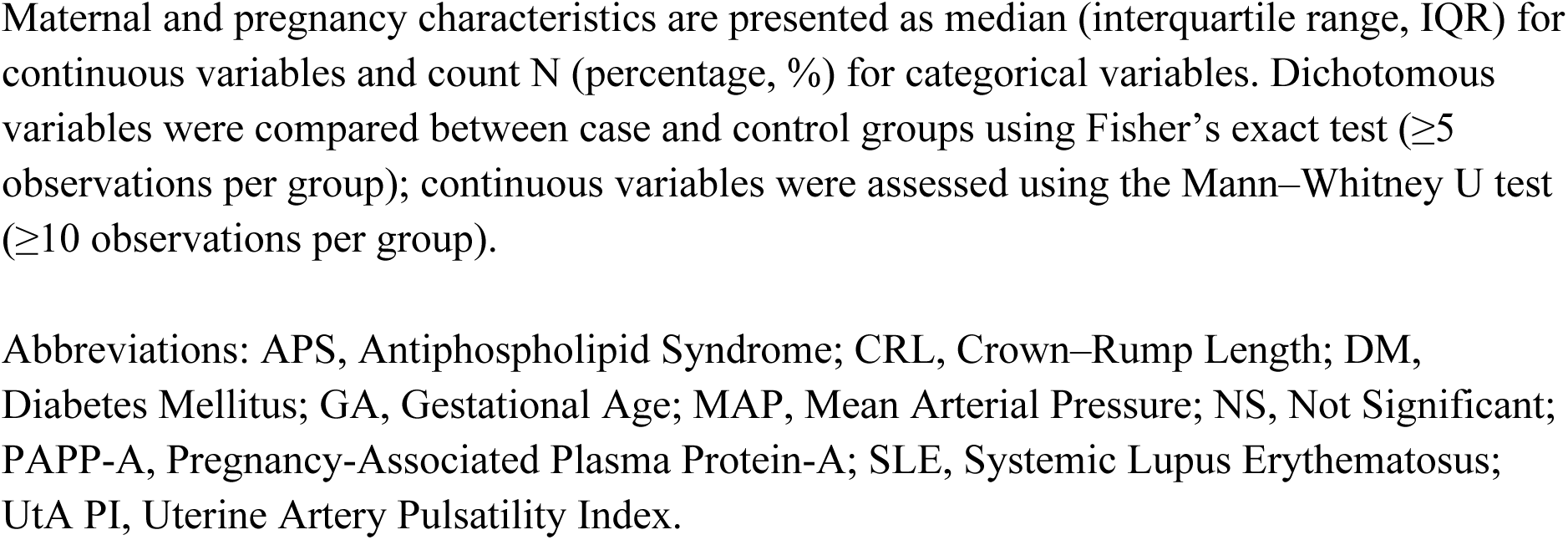
Cohort characteristics.

Control pregnancies resulted in delivery of phenotypically normal neonates at term with birthweights appropriate for gestational age; none of the control participants had a history of chronic hypertension. Compared with controls, both preterm and term preeclampsia groups showed significant differences in first-trimester risk factors, including mean arterial pressure (MAP; p < 0.001 for both), and a higher proportion of participants self-identifying as Black ethnicity (preterm: p < 0.01; term: p < 0.05).

Differences specific to the preterm preeclampsia group included elevated uterine artery pulsatility index (UtA-PI; p < 0.01). Maternal age and weight were modestly higher in the term preeclampsia group compared with controls (both p < 0.05), while gestational age at sampling did not differ between groups.

Perinatal outcomes differed markedly across cohorts. Preterm preeclampsia was associated with significantly earlier delivery and lower birthweight than controls (both p < 0.001). Term preeclampsia pregnancies delivered earlier and had lower birthweights (p < 0.05 and p < 0.01, respectively).

No other comparisons were made for groups with less than five observations for dichotomous variables and ten observations for continuous variables due to a lack of statistical power.

### Sample Processing

The optimized cfDNA extraction workflow increased cfDNA recovery from patient plasma relative to the QIAamp MinElute ccfDNA Midi Kit, with a median ∼1.7-fold increase in total yield across matched samples (paired t-test, p < 0.0001). Recovery of longer cfDNA fragments also increased (∼2.5-fold). Libraries generated using this workflow produced a median of 89.09 Gb raw and 69.84 Gb aligned data per sample, exceeding PromethION vendor benchmarks for cfDNA (supplementary data: Results – Sample Processing).

### Multi-Modal Preeclampsia Prediction

We developed a multi-modal classifier to predict preterm and term preeclampsia from first-trimester cfDNA by integrating fragmentomic, epigenetic, and clinical signals. Because no single modality fully separated cases from non-cases, we used an ensemble strategy combining multiple feature spaces.

Individual component models achieved AUROCs of 0.60 to 0.70, indicating limited discriminatory capacity alone. The ensemble approach improved performance by ∼20 AUROC percentage points for both disease subtypes. Final ensemble classifiers achieved AUROC (95% CI) values of 0.85 (0.77-0.91) for preterm preeclampsia and 0.84 (0.76-0.91) for term preeclampsia (Figure 3), demonstrating improved predictive accuracy compared with any single feature class.

**Figure 3:**
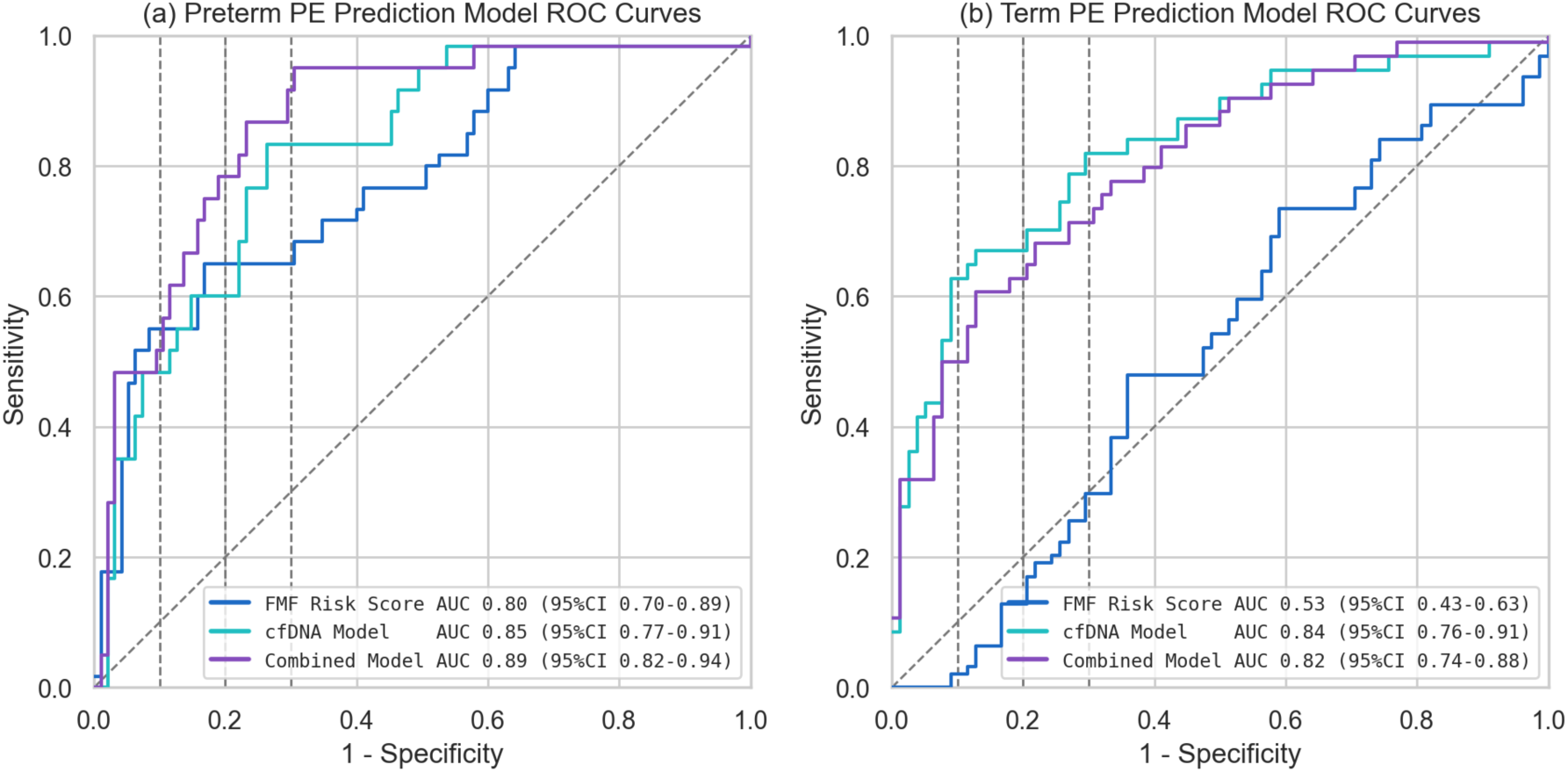
Predictive performance of multi-modal cfDNA models for preterm and term preeclampsia. Receiver operating characteristic (ROC) curves comparing the performance of the Fetal Medicine Foundation (FMF) first-trimester risk score (blue), the cfDNA-based multi-modal model (turquoise), and the combined model integrating cfDNA features with the FMF risk score (purple). (a) ROC curves for prediction of preterm preeclampsia. (b) ROC curves for prediction of term preeclampsia. The area under the ROC curve is reported for each model, with corresponding 95% confidence intervals.

The FMF score calculated based on parameters shown in Table 1 (excluding PlGF) achieved AUROC (95% CI) values of 0.80 (0.70-0.89) for preterm preeclampsia and 0.53 (0.43-0.63) for term preeclampsia. The combined model improved AUROC by ∼9 percentage points for preterm disease and ∼29 percentage points for term disease relative to the FMF score alone (Figure 3).

The multi-modal cfDNA classifier showed higher sensitivity than the FMF score at 70% and 80% specificity (Table 2). At 90% specificity, the FMF score retained a modest sensitivity advantage (∼6 percentage points), consistent with its design. In contrast, cfDNA-derived signals showed a more continuous risk distribution, providing more balanced performance across clinically relevant specificity ranges, particularly at 70-80%.

**Table 2:**
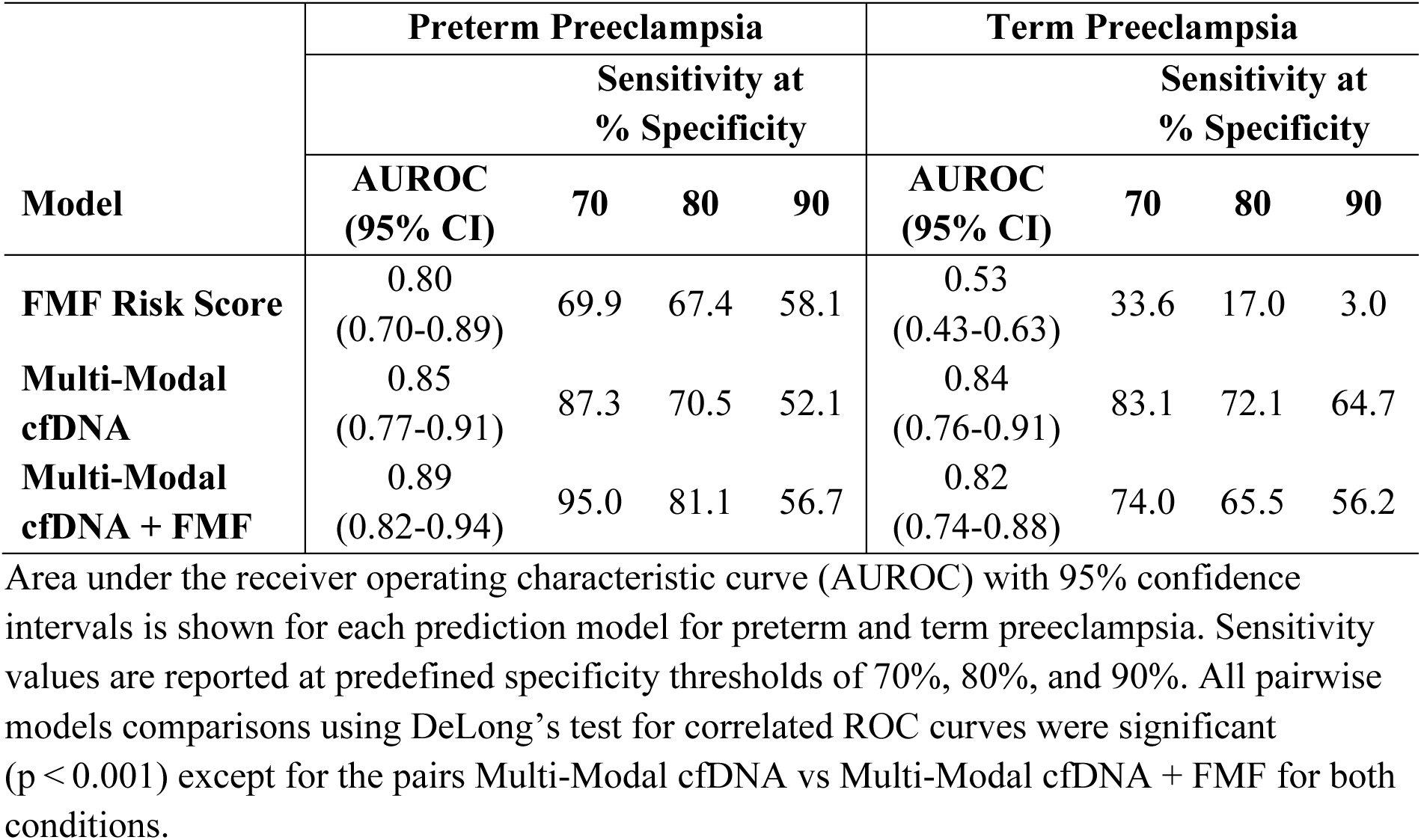
Discriminatory performance of FMF, cfDNA-based, and combined models for preterm and term preeclampsia.

Subgroup analysis stratified by baseline risk using the FMF first-trimester risk score (low vs high risk threshold 1 in 100) assessed robustness across risk strata. Within the cohort, FMF classified 7 of 30 preterm and 18 of 47 term preeclampsia cases as low risk, indicating that 23.3% and 38.3% of cases would not have been identified by FMF-based screening alone.

The cfDNA-based model maintained discriminative performance across baseline risk strata and identified additional cases not captured by FMF risk stratification (2 preterm and 13 term), while preserving specificity and achieving even higher specificity in high-risk pregnancies.

### Simulated Population-Level Screening Performance

In the simulated real-world screening cohort, the positive likelihood ratio was 4.25 for preterm preeclampsia and 3.83 for term preeclampsia. For preterm disease, this exceeded the FMF score (positive likelihood ratio 3.72). The negative likelihood ratio for preterm preeclampsia was 0.214, lower than the FMF value of 0.325, indicating greater residual risk reduction after a negative cfDNA-based test.

Using these likelihood ratios, the negative predictive value for preterm preeclampsia was 99.5% and the positive predictive value 9.8%. For term preeclampsia, the negative and positive predictive values were 97.3% and 23.7%, respectively (Table 3). These results reflect the low population prevalence of both conditions while highlighting strong rule-out performance of the cfDNA-based classifier, particularly for placental-associated preterm disease.

**Table 3:** Simulated population-level screening performance for preterm and term preeclampsia.

|  | **Preterm**<br>**Preeclampsia** | **Term**<br>**Preeclampsia** |
| --- | --- | --- |
| **Prevalence** | 2.5% | 7.5% |
| **Accuracy** | 80.6% | 80.4% |
| **Sensitivity** | 82.8% | 72.5% |
| **Specificity** | 80.5% | 81.1% |
| **PPV** | 9.8% | 23.7% |
| **NPV** | 99.5% | 97.3% |
| **Screen Positive Rate** | 21.1% | 23.0% |
| **Pre-test Prob. (Odds)** | 2.5% (0.026) | 7.5% (0.081) |
| **Post-test +Prob. (Odds)** | 9.8% (0.109) | 23.7% (0.311) |
| **Post-test –Prob. (Odds)** | 0.5% (0.005) | 2.7% (0.027) |
| **Likelihood Ratio+** | 4.249 | 3.830 |
| **Likelihood Ratio–** | 0.214 | 0.339 |

Reported metrics include accuracy, sensitivity, specificity, positive and negative predictive values (PPV, NPV), screen-positive rate, and corresponding pre- and post-test probabilities and odds. Likelihood ratios for positive and negative test results are shown to quantify the change from pre-test to post-test probability.

### Biological Motivation

Incorporation of tissue-resolved features significantly improved classification performance in a subtype-specific manner. For preterm preeclampsia, read-level fragmentomic features – particularly fragment end motifs – contributed significantly to predictive performance (McNemar’s test, p < 0.05), consistent with a placental-driven disease process. In contrast, for term preeclampsia, nucleosomal accessibility profiles were more informative when derived from maternal resolved cfDNA features than from unresolved signals (McNemar’s test, p < 0.01). These findings indicate that the cfDNA classifier captures biologically meaningful, tissue-specific signals distinguishing placental-associated preterm from immune-associated term disease.

Ranked feature importances across ensemble components revealed minimal or weak negative correlation between preterm and term preeclampsia models (Spearman correlation, p < 0.0001; Figure 4), indicating that the classifiers learned distinct signal combinations per subtype, supporting hypothesis that subtypes reflect partially divergent biological processes detectable as early as the first trimester.

**Figure 4:**
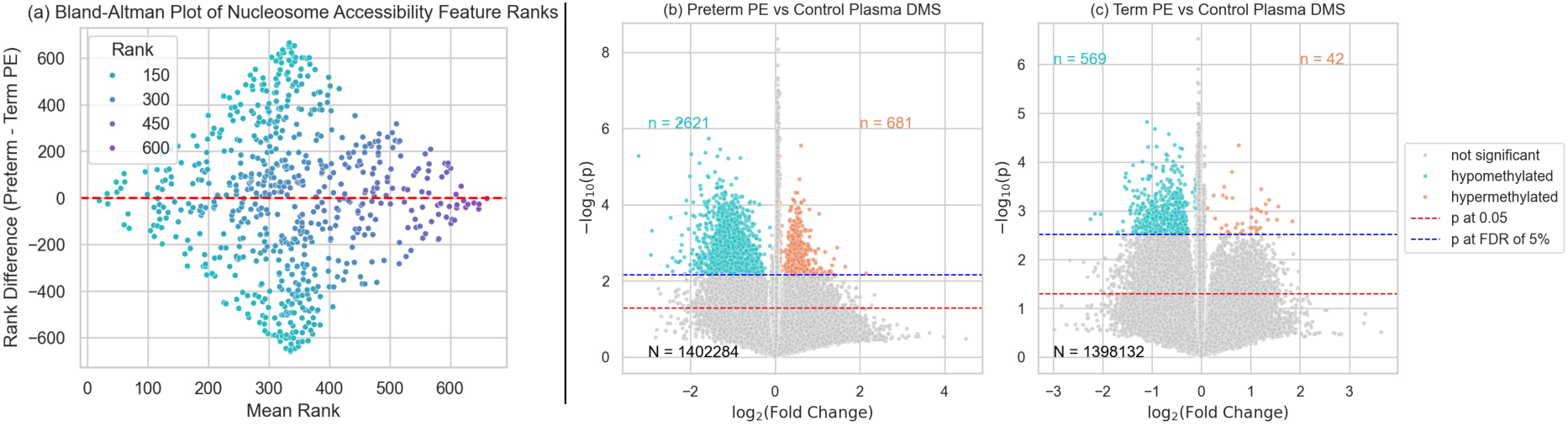
Subtype-specific feature learning and differential cfDNA methylation patterns. (a) Bland-Altman plot comparing ranked nucleosome accessibility feature importances derived from the preterm and term preeclampsia classifiers, shown for an illustrative feature modality. The absence of systematic agreement and the wide dispersion around zero indicate that nucleosome accessibility features are weighted differently between models, consistent with subtype-specific signal learning. (b) Volcano plot showing differentially methylated segments (DMSs) in first-trimester plasma cfDNA comparing preterm preeclampsia cases versus controls. (c) Volcano plot showing DMSs comparing term preeclampsia cases versus controls. DMSs passing a false discovery rate (FDR) threshold of 5% are highlighted, with hypomethylated segments shown in turquoise and hypermethylated segments in orange. The horizontal blue line indicates the FDR-adjusted significance threshold.

Functional enrichment analysis of differentially methylated segments in plasma identified 2,621 hypomethylated and 681 hypermethylated segments for preterm preeclampsia among ∼1.4 million candidate DMS, compared with 569 hypomethylated and 42 hypermethylated segments for term preeclampsia, consistent with a broader methylation signal in preterm disease.

For preterm preeclampsia, enrichment analysis highlighted angiogenesis and placental development pathways, including the RHO GTPase cycle (FDR < 0.01) and upstream Signaling by Receptor Tyrosine Kinases (FDR < 0.05). Pathways involving cadherin-11 expression and function were also enriched, implicating calcium-dependent cell-cell adhesion mechanisms required for trophoblast invasion and placental morphogenesis.^25^

Term preeclampsia showed fewer enriched pathways overall. Most prominently, pathways related to gastrulation and early developmental patterning were overrepresented (FDR < 0.05). Although gastrulation is completed early in embryogenesis, these pathways include regulatory programs reused in placental differentiation and maternal-fetal interface remodeling.^26^

## Comment

### Principal Findings

This study demonstrates the feasibility of predicting both preterm and term preeclampsia from first trimester cfDNA derived from routine prenatal plasma samples. By integrating fragmentomic and methylation-derived signals with tissue-resolved features and maternal clinical characteristics, the multi-modal classifier extends prior cfDNA approaches that have largely focused on preterm preeclampsia or single molecular modalities.^19–22^

To our knowledge, these findings provide the first evidence that cfDNA captures biologically relevant information across the full clinical spectrum of preeclampsia, including term disease, from a routine first-trimester blood sample. These results suggest potential for scalable population-level screening and risk-adapted prevention and management strategies.

### Results in the Context of What is Known

cfDNA fragmentation profiles reflect cell-type-specific nucleosome positioning and chromatin accessibility.^21^ The predictive contribution of fragmentomic features observed is consistent with early placental dysfunction and maternal endothelial or immune perturbations leaving detectable cfDNA signatures before clinical onset.

DNA methylation provides a complementary modality capturing stable, cell-type-specific regulatory states.^16,27^ The predictive value of methylation-derived features supports early epigenetic alterations associated with abnormal placentation or maladaptive maternal responses. Compared with univariate measures such as fetal fraction, methylation enables tissue-specific resolution relevant to heterogeneous phenotypes such as term preeclampsia.

Tissue-resolved cfDNA features allow attribution of signals to maternal or placental origins, helping resolve the differing pathophysiologies of preterm and term preeclampsia and supporting the view of preeclampsia as a biologically heterogeneous disorder.^28^ Improved molecular subtyping may ultimately inform targeted preeclampsia therapies beyond current approaches such as low-dose aspirin.

Functional enrichment analysis provides biological context for the subtype-specific signals captured by the classifiers. For preterm preeclampsia, enrichment of RHO GTPase signaling, receptor tyrosine kinase pathways, and cadherin-mediated adhesion is consistent with impaired placentation, including disrupted trophoblast invasion, angiogenesis, and endothelial function.^29^ In contrast, term preeclampsia showed fewer enriched pathways, including developmental patterning programs, consistent with a more heterogeneous and less placenta-centric pathophysiology.^19^ Together, these observations support the biological plausibility of the multi-modal cfDNA framework and the integration of fragmentomic, methylation, and tissue-resolved signals for early subtype-specific risk prediction.

### Clinical Implications

All cfDNA features were derived from plasma samples and sequencing data compatible with standard first-trimester NIPT workflows, allowing screening from a single blood draw. This approach could facilitate early risk stratification and address the persistent quality gap in preeclampsia prevention identified by professional societies.^30^

Integration with existing NIPT workflows may be particularly relevant for term preeclampsia, for which no effective first-trimester screening approach currently exists. Prospective validation in larger and unselected populations will be required to confirm performance estimates and evaluate clinical utility, including in pregnancies complicated by gestational hypertension and gestational diabetes.

### Research Implications

The FMF algorithm was designed to identify early-onset and preterm preeclampsia and relies heavily on clinical and hemodynamic markers that perform well in high-risk populations.^8,31^ It was not intended to predict term disease, which is more heterogeneous and less tightly linked to early placental insufficiency. The complementary performance profiles observed here should therefore be interpreted considering these differing objectives rather than as a direct competition between methods.

Further refinement of tissue-resolved modeling may enable improved biological disease subtyping and provide insight into early mechanisms underlying maternal and fetal complications beyond preeclampsia.

### Strengths and Limitations

This study represents the largest cfDNA-based preeclampsia prediction analysis from routine first-trimester blood samples in a heterogenous multi-ancestry cohort and the first to apply tissue-resolved features derived from single-molecule sequencing. The availability of matched maternal and placental tissue enabled biological anchoring of cfDNA signals, strengthening tissue-of-origin inference and interpretability.

A technical strength is the multi-modal strategy that preserves native cfDNA fragment lengths and endogenous methylation on the same molecules. Unlike short-read, amplification-based workflows, this approach enables integrated fragmentomic and epigenetic analysis without PCR or conversion-induced bias.

The primary limitation is cohort size, which restricts subgroup analyses and requires cautious interpretation of performance estimates. Larger prospective validation studies are therefore required and are currently in planning.

### Conclusions

First-trimester cfDNA sequencing enables prediction of both preterm and term preeclampsia by capturing molecular signals reflecting placental and maternal contributions across the disease spectrum. A full-spectrum early screening test could support earlier risk stratification and inform prevention, surveillance and timing-of-birth strategies, particularly for term disease.

By providing an objective molecular risk assessment early in pregnancy, this approach addresses the quality gap highlighted by the Society for Maternal-Fetal Medicine, namely the under identification of at-risk pregnancies and the suboptimal uptake of evidence-based preventive interventions for preterm disease.

Compatibility with routine NIPT workflows provides a practical and scalable pathway for combined fetal and maternal screening, positioning cfDNA-based approaches as a core component of next-generation prenatal care.

## Data Availability

All data produced in the present study are available upon reasonable request to the authors.

