## Supplemental Data for "First-Trimester Multi-modal cfDNA Analysis for Prediction of Preterm and Term Preeclampsia"

### Supplementary Materials

#### DNA Extraction and Sequencing

##### Cell-free DNA Extraction and Sequencing

Cell-free DNA (cfDNA) was extracted from maternal plasma using the aitios® cfDNA internal standard operating protocol v1.

cfDNA quality and fragment size distribution were assessed using the QIAxcel High Sensitivity assay. Samples passing quality control were purified using AMPure XP beads and quantified by fluorometry. Libraries were prepared from purified cfDNA for long-read sequencing using the Oxford Nanopore Technologies (ONT) ligation-based protocol (Ligation Sequencing Kit V14) with R10.4.1 flow cells.

Prior to adapter ligation, cfDNA underwent DNA repair and end-preparation using NEBNext FFPE DNA Repair Mix and Ultra II End Prep reagents, with incubation conditions optimized for fragmented and low-input DNA. Cleanup steps employed modified bead-to-sample ratios and extended binding and elution times to improve recovery of short and long cfDNA fragments. Sequencing adapters were ligated using Quick T4 DNA ligase, followed by bead-based purification and library quantification.

Sequencing was performed on a PromethION 24 instrument using R10.4.1 flow cells. Libraries were primed and loaded according to the manufacturer's recommendations. Runs were configured using Dorado v.1.1.0 with super-accurate canonical (v5.2.0) and modified basecalling (v5.2.0, 5mCG, 5hmCG, v1), generating BAM-formatted output for downstream analysis.

##### Genomic DNA Extraction and Sequencing

Genomic DNA (gDNA) was extracted from reference tissues using the aitios® gDNA internal standard operating protocol (SOP) v1.

High-molecular-weight gDNA was sheared to an average fragment length of approximately 10 kb using g-TUBEs and size-selected to remove fragments shorter than 5 kb. DNA quantity and purity were assessed spectrophotometrically, and fragment integrity was verified using QIAxcel DNA Screening Kit.

gDNA libraries were prepared for long-read sequencing using the Oxford Nanopore Technologies (ONT) ligation-based protocol (Ligation Sequencing Kit V14) with R10.4.1 flow cells. Input DNA (1-3 μg) underwent DNA repair and end preparation using NEBNext FFPE DNA Repair and Ultra II End Repair/dA-Tailing reagents to correct base damage, nicks, and blocked ends and to generate 5' phosphorylated, 3' dA-tailed fragments. Cleanup steps were performed using AMPure XP beads, with bead-to-sample ratios adjusted according to DNA input amount.

Adapter ligation was performed according to the manufacturer's recommendations. Fragment size retention during cleanup was adjusted according to sample type: Long Fragment Buffer was used to enrich for fragments >3 kb where sufficient input material was available, while Short Fragment Buffer was used for low-input placental villi samples to retain the full fragment size distribution.

Final libraries were eluted in elution buffer, quantified by fluorometry, and sequenced using the same platform and run configuration as the cfDNA libraries.

Sequencing was performed on a PromethION 24 as described above.

#### Bioinformatics Processing

Sequencing data derived from both cfDNA and reference gDNA samples were processed using a unified bioinformatics pipeline (Nextflow v≥25.04). Reads were aligned to the T2T-CHM13v2 reference genome using minimap2 v2.28[1] with ONT-optimized parameters (map-ont preset), retaining reads with a minimum read quality score of 10 and alignments with a minimum mapping quality of 10. Read-level and genome CpG-site methylation levels were derived using modkit v0.4.1[2]. Genome-wide coverages were computed using mosdepth v0.3.10[3]; alignment flag distributions were assessed using samtools v1.21[4] and additional sequencing and alignment quality metrics were compiled using NanoPlot v1.46[5].

##### Germine Variant Calling

Germline short variants, including single-nucleotide variants (SNVs) and small insertions and deletions (indels), were detected using Clair3 v1.0.10, parameterized with the ONT R10.4.1 super-accurate model v500. Variant calling was performed against the T2T-CHM13v2 reference genome. Resulting variant call format files were sorted and normalized using bcftools v1.21, including left-alignment and decomposition of multi-allelic sites (bcftools norm -multiallelics). Variants with genotype quality scores below 20 were excluded from downstream analyses.

##### Quality Control

Sample quality was assessed using a two-stage quality control (QC) workflow combining multivariate sequencing and alignment metrics with sample identity verification. Primary QC focused on sequencing- and alignment-derived characteristics and was applied to both cfDNA and reference gDNA data.

Sequencing summary statistics and alignment metrics were jointly evaluated using principal component analysis. Multivariate outliers were identified using Hotelling's $T^2$ statistic, with a 5% false-positive threshold. Samples flagged as outliers were further examined by manual inspection of individual quality metrics. Samples with mean sequencing coverage below 10x were excluded from downstream analyses. In addition, cfDNA plasma samples showing evidence of genomic DNA contamination, inferred from aberrant fragment length distributions, were removed.

To verify sample identity and detect potential sample mix-ups, a germline SNV-based concordance analysis was performed for pregnancies with matched cfDNA, buffy coat, and placental villi samples. This analysis assessed genetic concordance within and between subjects to identify inconsistencies indicative of mislabeling or cross-contamination.

For each pregnancy, pairwise comparisons were performed between all available sample types (plasma-buffy coat, plasma-placenta, and buffy coat-placenta). For each pairwise comparison, SNV concordance was quantified using sensitivity, defined as the proportion of SNVs detected in one sample that were also detected in the corresponding paired sample. Sensitivity was computed in both directions for each pairwise comparison, yielding up to six concordance values per subject.

Cohort-level distributions of sensitivity values were generated separately for each comparison type. Samples exhibiting sensitivity values falling outside the expected distribution for their comparison group were flagged as potential outliers. Low within-subject concordance was interpreted as evidence of sample degradation, insufficient sequencing quality, or incorrect sample assignment.

To identify potential sample mix-ups or cross-contamination between pregnancies, SNV concordance was also assessed between samples derived from different subjects. Each sample type from a given pregnancy was compared against all sample types from every other pregnancy in the cohort. Sensitivity values were computed as described above.

Between-subject comparisons exhibiting unusually high concordance relative to the cohort-level distribution were flagged as potential mislabeling or cross-contamination events, as unrelated pregnancies are not expected to share high levels of germline SNV concordance.

**cfDNA Fragmentomics**

Fragmentomic analyses were performed exclusively on cfDNA sequencing data. For each read, fragment length, 5’ and 3’ end sequence motifs, and terminal nucleotides were characterized. End motifs were defined as 4-mers at both fragment termini; in cases of incomplete adapter trimming, soft-clipped segments were used. Fragment end nucleotide composition was derived from the 5’ and 3’ terminal nucleotides.

Fragment length distributions were summarized using three bins (≤150 bp, 151–500 bp, and >500 bp). End-motif frequencies were stratified into short (≤500 bp) and long (>500 bp) fragment categories. End-nucleotide frequencies were computed across predefined fragment length intervals, using 27 bins of variable size tailored separately for 5’ and 3’ fragment ends. In addition, the mean, median, and standard deviation of fragment length were calculated for each sample.

For each sample, count-based fragmentomic features were normalized independently within five disjoint feature groups - fragment length distributions, 5’ end motifs, 3’ end motifs, 5’ end nucleotide-by-length distributions, and 3’ end nucleotide-by-length distributions - such that feature values within each group summed to unity to enable direct comparison of relative fragmentation patterns across samples.

Nucleosome accessibility features were computed using a custom Python implementation based on previously described methodologies[6,7], adapted for long-read sequencing data. Genomic regions of interest were defined using the CATlas database[8] of single-cell ATAC-seq profiles, comprising cell-type–specific open chromatin regions. For each cell type represented in CATlas, the top 10,000 regions ranked by peak signal were selected. Sequencing coverage was computed in fixed windows centered on these regions, corrected for GC-related coverage biases, and aggregated by cell type.

Three nucleosome accessibility metrics were derived: mean coverage, defined as the average midpoint coverage within a ±1,000 bp window; central coverage, defined as the average midpoint coverage within a ±30 bp window; and an oscillatory amplitude metric derived from the smoothed fast Fourier transform of the midpoint coverage signal across a ±960 bp window.

#### Reference Tissue Differential Methylation

Differentially methylated segments (DMSs) were identified independently in placental villi and maternal buffy coat reference tissues for each classification task: preterm preeclampsia versus all non-preterm samples, and term preeclampsia versus all non-term samples. DMS discovery was based on the method by Loyer et al.[9] For each tissue and comparison, candidate DMSs were first filtered using a nominal p-value threshold of 0.05 and subsequently ranked using a composite score that integrated statistical significance and effect size.

The composite score combined the (negative log-transformed) p-value with a weighted contribution of (i) the absolute $\log_2$ fold-change of class-specific mean methylation levels and (ii) the absolute difference in methylation between classes. Both effect size components were z-score normalized prior to combination, and equal weighting was applied to balance their contributions. The p-value term was dampened using a concave transformation to reduce dominance of extremely small p-values. For each tissue and comparison, the top 2,500 ranked DMSs were retained as loci of interest.

#### Fetal Fraction Estimation

An origin atlas of DMSs was constructed for tissue deconvolution and fetal fraction estimation. For each tissue class, the top 250 hypomethylated and hypermethylated segments were selected based on effect size, requiring a multiple-testing–corrected p-value <0.05 and at least three CpG sites per segment.

For plasma cfDNA samples, segment-level methylation values were projected onto the origin atlas and weighted by the square root of total signal counts per segment. Tissue contributions were estimated using non-negative least squares with iterative Huber weighting to improve robustness. Estimated coefficients were normalized to sum to unity, yielding an estimate of the fetal fraction as the placental contribution.

#### Multi-Modal Read Classification

Multi-modal cfDNA read classification was performed using a probabilistic framework integrating fragmentomic and epigenetic features to assign individual cfDNA fragments to maternal or placental tissue of origin. The classifier was trained and evaluated using a large ground truth dataset derived from matched maternal buffy coat, placental villi, and plasma samples, in which cfDNA read origin was established via germline SNVs. This dataset represents one of the largest SNV-defined cfDNA read-level ground truth resources reported to date in the prenatal setting.

The classifier enabled tissue-resolved feature extraction, fetal fraction estimation, and downstream modeling used in the present study. A comprehensive description of the read classification framework, reference profile construction, statistical formulation, and performance evaluation is beyond the scope of this manuscript and will be reported separately in a dedicated methodological publication currently in preparation.

#### Multi-Modal Preeclampsia Prediction

The multi-modal, tissue-resolved preeclampsia classifier infers preterm and term preeclampsia risk from cfDNA extracted from plasma. Risk prediction models were constructed using fragmentomic and epigenetic features as well as clinical and demographic features.

Fragmentomic features included fragment length distributions, 5′ and 3′ end-motif frequencies, fragment end nucleotide compositions, and global fragment length statistics; nucleosome accessibility features, comprising mean coverage, central coverage, and oscillatory amplitude; tissue-resolved fragmentomic and nucleosome features, computed separately on cfDNA reads classified as placental-derived and maternal (buffy coat)-derived using identical processing pipelines. As a result, fragmentomic and nucleosome accessibility features were expanded three-fold (global, placental-specific, and maternal-specific). For fragmentomic features, additional log fold-change representations between placental and maternal components were included.

Epigenetic features included plasma cfDNA methylation signals that were projected onto placental- and maternal-derived differentially methylated segments (DMS), generating per-segment methylation features that summarize disease-associated epigenetic differences at loci defined from reference tissues.

Clinical and demographic features included maternal age, weight, height, gestational age at sampling, self-reported ethnicity, smoking status, medical history (chronic hypertension, diabetes mellitus type 1 or 2, systemic lupus erythematosus, antiphospholipid syndrome), family history of preeclampsia, method of conception, parity, and arterial blood pressure measurements; fetal parameters, including fetal fraction and fetal sex derived from cfDNA.

Features with more than 10% missing observations or with ≥95% invariant values were removed prior to normalization. Missing values were imputed during model training as part of the preprocessing pipeline.

**Model Development**

Model training followed a modular ensemble framework. Three learning algorithms-ElasticNet regression, stochastic gradient descent classifiers, and gradient-boosted decision trees-were trained independently on feature subsets grouped by feature type. Hyperparameter optimization was performed using stratified 10-fold cross-validation. Preprocessing steps, including feature scaling (unit-variance or interquartile-range scaling), mean imputation of missing values, and class imbalance correction (random up-sampling, synthetic minority over-sampling, or no resampling), were incorporated into the hyperparameter optimization process.

The resulting component models were combined into a soft-voting ensemble classifier, assigning a class probability to a sample by averaging across class probabilities from all component models. Final reported class probabilities for all samples were obtained by concatenating predictions on validation splits of a 10x cross-validation on the full dataset using optimized hyperparameters.

**Functional Enrichment Analysis**

To explore the biological processes represented in plasma cfDNA methylation changes, functional enrichment analysis was performed on DMSs identified between case and control samples using a similar methodology as for the reference tissues (supplementary materials: Reference Tissue Differential Methylation). Given the limited interpretability of machine-learning features derived from fragmentomic and nucleosome-based summary statistics, enrichment analysis was restricted to locus-resolved methylation changes rather than model-derived feature weights.

DMS were identified from plasma cfDNA methylation profiles and filtered to retain CpG segments comprising at least five CpG sites, in line with evidence that DNA methylation changes tend to occur in coordinated blocks rather than isolated CpGs.[10] DMS were considered significant if they met a false discovery rate (FDR) threshold of 5% and exhibited an absolute methylation difference (log-fold change) of at least 0.1 between case and control samples, to reduce noise from small methylation changes.

Gene annotations corresponding to significant DMS loci were generated using an in-house R pipeline. Pathway enrichment analysis was subsequently performed using the PANTHER knowledgebase[11] and the Reactome Pathway Database[12] with default parameters. Analyses were conducted separately for preterm and term preeclampsia to assess subtype-specific biological signals.

#### Results – Sample Processing

cfDNA and gDNA were processed using laboratory workflows optimized to maximize sequencing yield and, for cfDNA, to ensure efficient recovery of native cfDNA fragment length distributions.

To evaluate extraction performance, comparative experiments were performed on patient plasma samples, with bulk plasma (CLINIQA) included as an independent control. cfDNA was extracted either using a custom cfDNA extraction workflow or the QIAamp cfDNA kit (Qiagen), which served as a state-of-the-art comparator.

cfDNA yield was quantified using both Quantus fluorometry and qPCR to ensure consistent quantification across measurement modalities. Across matched samples, the custom extraction workflow produced significantly higher cfDNA amounts than the QIAamp cfDNA kit (paired t-test, $p < 0.0001$), with a median increase in total cfDNA yield of approximately 1.7-fold (Figure S1).

Recovery of longer cfDNA fragments was evaluated using QIAxcel electropherogram analysis. Custom extraction resulted in a median 2.5-fold increase in the abundance of long cfDNA fragments (>500 bp) compared with QIAamp-based extraction.

Sequencing libraries were prepared from extracted cfDNA and sequenced on PromethION flow cells. Libraries generated using the optimized workflow produced a median of 89.09 Gb (IQR 82.51-94.43 Gb) of raw sequencing data per sample, corresponding to a median of 69.84 Gb (IQR 60.64-79.22 Gb) of aligned data. These yields exceeded vendor-reported performance benchmarks for cfDNA sequencing on a PromethION flow cell (66 Gb raw output and 50 Gb aligned output), indicating robust sequencing performance using the optimized workflow.

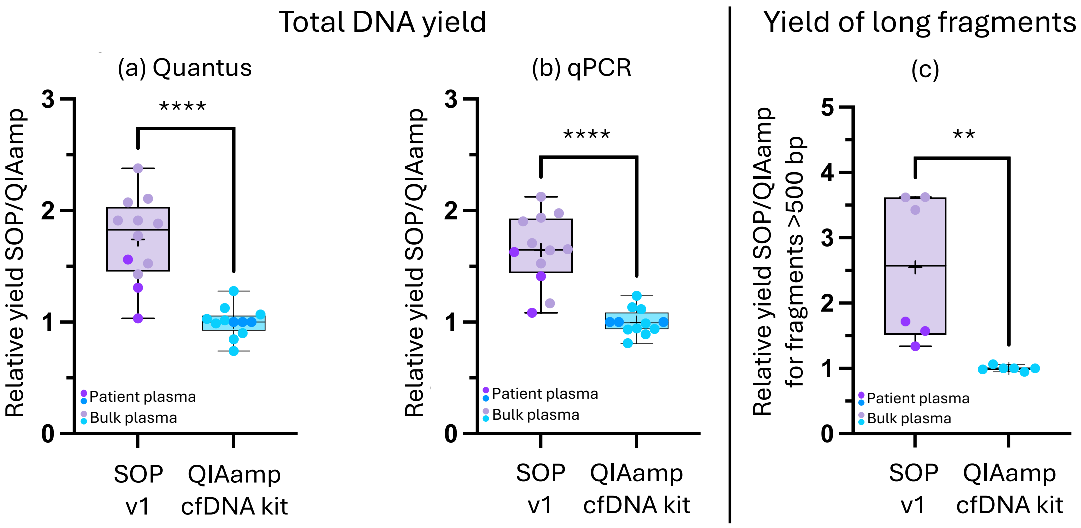


Figure S1: Boxplots comparing the relative cfDNA yield obtained with custom extraction versus the QIAamp cfDNA kit across matched plasma (n=3) and bulk samples (n=9): relative yield measured with (a) Quantus fluorometry and (b) qPCR, demonstrating a consistent increase in overall cfDNA recovery. (c) Relative yield of long cfDNA fragments (>500 bp) based on electropherogram-derived fragment abundance, indicating preferential enrichment of long fragments with custom cfDNA extraction. ** denotes $p < 0.01$; **** denotes $p < 0.0001$.
